# Performance of suspected influenza case definition before and during the COVID-19 pandemic

**DOI:** 10.1101/2020.06.01.20119446

**Authors:** Efrén Murillo-Zamora, Carlos Hernandez-Suarez

## Abstract

**Background:** The influenza-related burden remains high and the COVID-19 pandemic may difficult its accurate surveillance. This study aimed to evaluate the performance, before and during the COVID-19 pandemic, of the case definition of suspected influenza used in community surveillance in Mexico. Methods: A cross-sectional analysis of a cohort study took place and cases fulfilling the suspected case criteria (*n* = 20, 511), and with laboratory-conclusive evidence (quantitative real-time polymerase chain reaction) to confirm or discard influenza virus infection, were analyzed.

**Results:** A high sensitivity and modest specificity was documented, and this later decreased during the COVID-19 outbreak, as well as its diagnostic accuracy. However, no significant differences were observed in the Area Under the Receiver Operating Characteristics among the analyzed periods.

**Conclusions:** The evaluated case definition remains to be a cost-effective alternative to identify patients who may benefit from influenza-specific antiviral drugs, even during the COVID-19 global outbreak.

## Background

The influenza-related burden remains high globally despite vaccination efforts [1]. In the northern hemisphere, seasonal cases start in October and tail off by May [2]. The case definitions of suspected influenza used in national and regional surveillance programs commonly differ from those recommended by WHO. In Mexico and according to normative standards [3], the case definition in patients aged 5 years or above includes the presence of fever (38 °C or higher), headache and cough accompanied by (at least 2): rhinorrhea, coryza, arthritis, arthralgia, myalgia, prostration, odynophagia, thoracic pain, abdominal pain, nasal congestion or diarrhea. Fever is not a cardinal symptom among elder subjects (65+ years old). Suspected cases fulfilling the criteria are classified as influenza-like illness (ILI) or severe acute respiratory infection (SARI) if systemic or distress symptoms are presented. This definition has several similarities to the proposed by the *Groupes Régionaux d’Observation de la Grippe* (*GROG*, the French acronym) [4], which has shown good performance in community surveillance of influenza [5].

The first registered cases of locally acquired coronavirus disease 2019 (COVID-19) in Mexico occurred in late February 2020 [6]. About three months later, over 75 thousand cases and 8 thousand deaths had been registered at national level [7]. Given that suspected cases of COVID-19 and influenza share clinical similarities [8], timely identification of these later, and which may benefit the start of neuraminidase inhibitors (NAIs) [9], may be challenging in source limited healthcare settings. We aimed to evaluate the performance of the influenza case definition before and during the COVID-19 pandemic in Mexico. We analyzed two consecutive flu seasons (2018–2020) for the benefit of a wider time framework.

## Methods

We conducted a cross-sectional analysis of a nationwide retrospective cohort study. Suspected cases of influenza among individuals aged 5 years or older, and registered during two consecutive seasons (2018–2020) in a normative system for the epidemiological surveillance of viral respiratory diseases (*SISVER*, the Spanish acronym), and which were later confirmed or discarded as cases of influenza virus infection, were eligible.

Quantitative real-time polymerase chain reaction (qRT-PCR; SuperScript® III Platinum® One-step RT-qPCR System) analyses were performed on clinical specimens (nasopharyngeal or deep nasal swab). A detailed description of laboratory methods employed in the *Mexican Institute of Social Security* (*IMSS*, the Spanish acronym) network was previously published [10].

The performance of case definition of suspected influenza case was evaluated in terms of sensitivity, specificity, accuracy, and positive and negative likelihood ratio (LR+/−). Age (5–9; 10–19; 20–44; 45–64 and 65 years or above) and time-stratified (according to symptoms onset: Oct. 2018-Feb. 2019; Mar. 2019-Apr. 2019; Oct. 2019-Feb. 2020; Mar. 2020-Apr. 2020) estimators were obtained. The fourth period was the pandemic one. The Area Under Receiver Operating Characteristic curves (AUROCs) and 95% confidence intervals (CI) was also computed. This study was approved by the Local Ethics in Health Research Committee (601) of the *IMSS* (R-2020-601-022).

## Results

Data from 20,511 cases were analyzed. The overall prevalence of laboratory-confirmed influenza in the study sample was 38.8% (*n* = 7, 955). Table 1 summarizes the estimates. The prevalence of laboratory-positive influenza was lower among elder subjects, particularly during the COVID-19 pandemic period (65+ years old, 13.2%). The mean sensitivity of case definition was high in all age groups and the last general estimate (92.7, 95% CI 91.3–94.1) was similar to the previous ones *p* = 0.274.

**Table 1.**
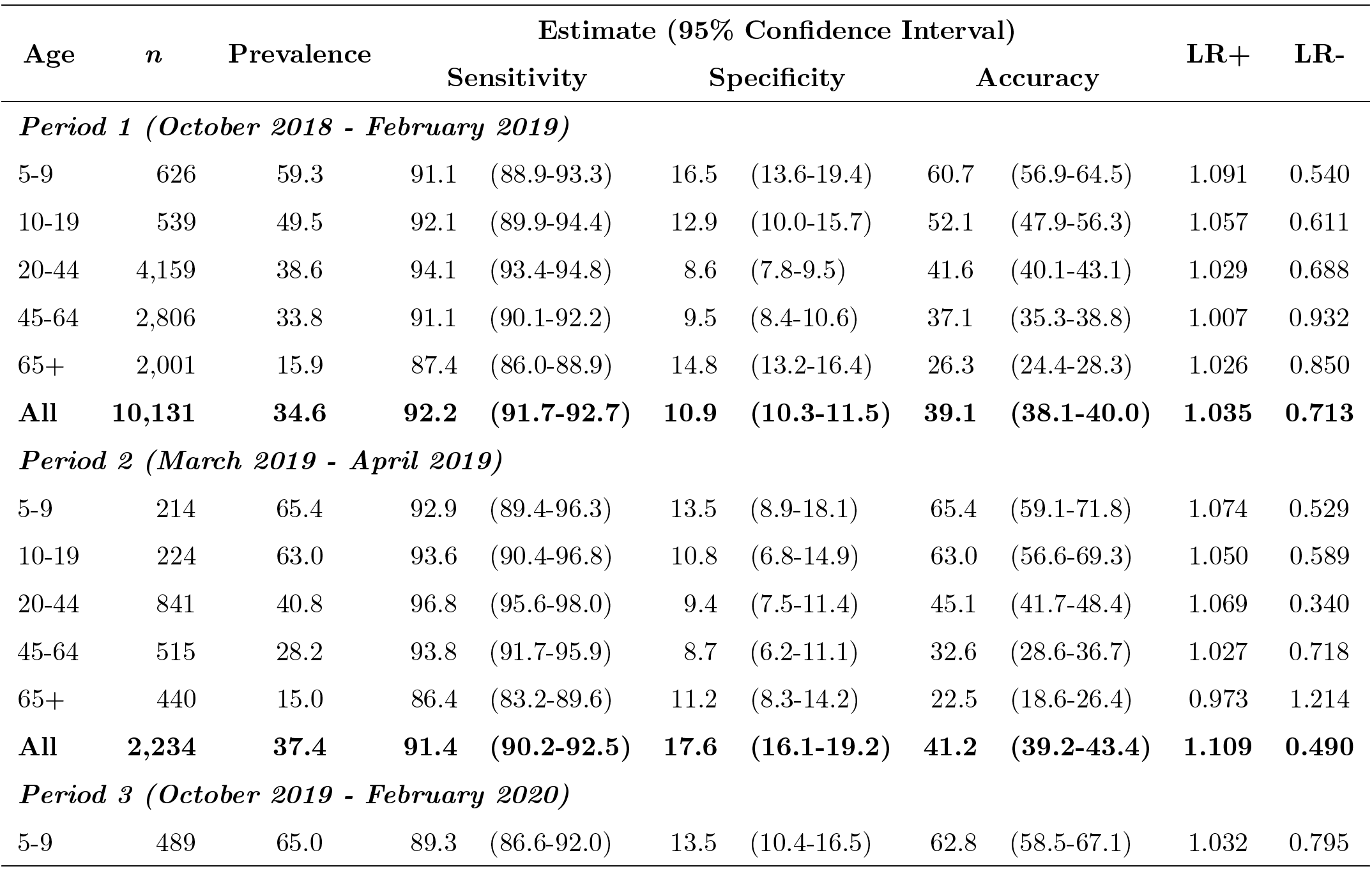
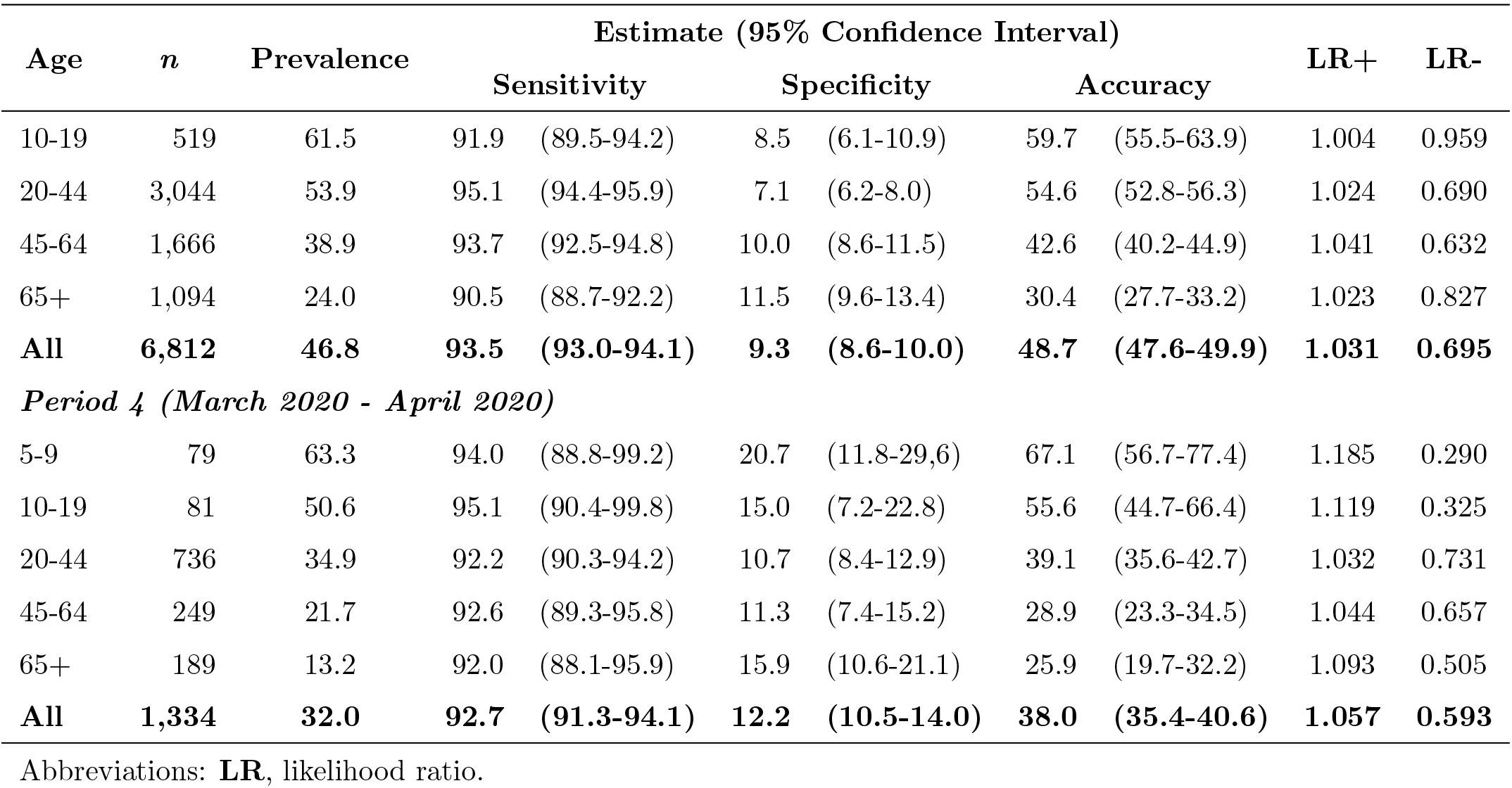
Performance of suspected influenza case definition, Mexico 2018–2020.

The overall specificity computed during the pandemic period was 12.2%(95% CI 10.5–14.0) and it was higher than the estimate from Period 3 (*p* = 0.001) but lower than the estimate from March-April 2019 (*p* < 0.001). The diagnostic accuracy went from 48.7% to 38.0% (22% decrease; *p* < 0.001) in Period 3 and 4, respectively; it was the similar to the accuracy from Period 1 (*p* = 0.459).

The AUROCs are presented in Figure 1 and ranged from 0.544 (95% CI 0.533–0.556) to 0.607 (95% CI 0.586–0.628). No significant differences were documented between the pre- and during-pandemic periods (*p* = 0.855).

**Figure 1:**
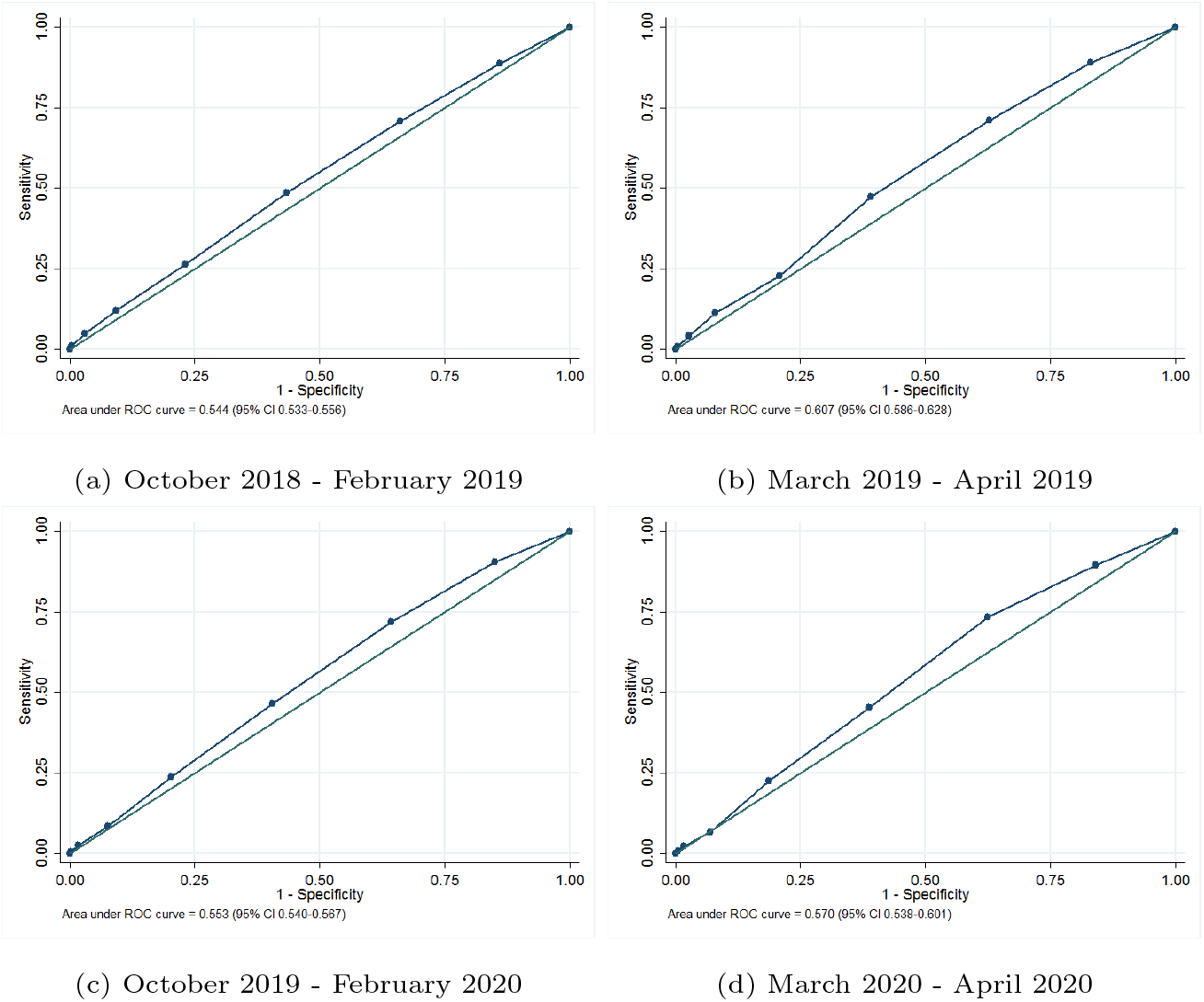
Area Under the Receiver Operating Characteristics (AUROC) and 95% Confidence intervals (CI) of suspected influenza case definition, Mexico 2018–2020

Note: No significant differences were documented between the pre- (a-c) and during-pandemic(d) periods (*p* = 0.855).

## Discussion

This study evaluated the performance of influenza case definition based on a national and normative influenza cohort. Our results suggest no significant changes in the evaluated parameters before and during the COVID-19 pandemic. The analyzed cohort study has several strengths and include: (i) influenza virus infection was confirmed by qRT-PCR analysis, which is the gold standard; (ii) the database included cases from all age groups and (iii) data of influenza A and B viruses was available.

Timely identification of influenza virus infection can assist healthcare providers in determining optimal strategies for preventing or treating influenza, including the use of antiviral drugs. These interventions also reduce the spread of influenza [11].

Multiple suspected influenza case definitions are currently being used worldwide and include, among others, the proposed by CDC [11], WHO [12], and the *GROG* [13]. All of them have a similar performance in detecting laboratory-positive cases and their sensitivity and specificity ranges from 90 to 96%, and from 7 to 21%, respectively [5]. The computed AUROCs by using any of these classifications are similar to those that were estimated in our study (*≈* 0.550).

No differences in the performance of the case definition or symptoms in influenza cases according to virus type were found in a recently published study [14]. The identified influenza virus sub-types were as following (*n* = 7, 955): A/H1N1, 57%; B/Victoria, 17%; A/H3, 16%; B/Yamagata, 9% and B unidentified, 1%.

Seasonal influenza vaccination has been proven to be cost-effective in the prevention of seasonal influenza [15], however low acceptance rates have been documented among Mexicans, even in high-risk groups (elderly, about 56%) [16]. Vaccination coverage among persons of productive age is even lower (20%) [17].

## Conclusions

Our findings suggest that the suspected case definition employed in community surveillance of influenza has a good performance, even during the COVID-19 pandemic. Therefore, this definition may be used in identifying patients who may benefit from early access to neuraminidase inhibitors. Timely use of antiviral drugs, together with immunization promoting, may reduce the social and economic burden of influenza.

## Data Availability

The datasets generated during and/or analysed during the current study are available from the corresponding author on reasonable request.

